# Representative Estimates of COVID-19 Infection Fatality Rates from Three Locations in India

**DOI:** 10.1101/2021.01.05.21249264

**Authors:** R. Cai, P. Novosad, V. Tandel, S. Asher, A. Malani

## Abstract

There are very few estimates of the age-specific infection fatality rate (IFR) of SARS-CoV-2 in low- and middle-income countries. India reports the second highest number of SARS-CoV-2 infections in the world. We estimate age-specific IFR using data from seroprevalence surveys in Mumbai (population 12 million) and Karnataka (population 61 million), and a random sample of economically distressed migrants in Bihar with mortality followup. Among men aged 50–89, IFR is 0.12% in Karnataka (95% C.I. 0.09%–0.15%), 0.53% in Mumbai (0.52%–0.54%), and 5.64% among migrants in Bihar (0–11.16%). IFR in India is approximately twice as high for men as for women, is heterogeneous across contexts, and rises much less at older ages than in comparable studies from high income countries.

---

Measurement of the infection fatality rate (IFR) for severe acute respiratory syndrome coronavirus 2 (SARS-CoV-2) has been a major objective for researchers since the beginning of the global pandemic (*1, 2*). Reliable estimates of the IFR are essential for policy decisions around non-pharmaceutical interventions and vaccine allocation plans (*1, 3*). In this paper, we estimate age-specific IFRs in three locations in India.

A range of estimates for age-specific SARS-CoV-2 IFRs now exists, the majority of which are based on data from high-income countries (*3*–*6*). Meta-analyses that estimate age-specific IFR in low- and middle-income countries (*7, 8*) rely on the assumption that crucial epidemiological characteristics (*e*.*g*. transmission dynamics, age-specific death rates) from high-income countries are generalizable to low-income settings.

Seroprevalence studies in low- and middle-income countries have given rise to several population-representative estimates of overall IFR (*9*–*14*). Ioannidis (*15*) additionally estimates IFR in several developing countries by combining population-representative seroprevalence surveys (*16*–*18*), and subpopulation-specific surveys (*e*.*g*. blood donors) (*13, 19*–*23*) with reported deaths. However, to date we are aware of only two studies in any LMIC that estimate age-specific IFRs, and these are limited to small or non-representative samples. The sample in Picon et al. (*24*) is representative for one city in Brazil; the sample in Verity et al. (*25*) is limited to repatriated international residents leaving Wuhan Province (China) in a two-day span. Thus, this paper is the first to provide representative age- and sex-specific IFR estimates for a large population (the combined population of Mumbai and Karnataka is 73 million) in a low- or middle-income setting.

Early models of lower-income settings assumed that age-specific infection mortality rates would be higher due to worse baseline population health and under-resourced healthcare systems (*8, 26, 27*). Other researchers observed low case fatality rates (CFR) in Sub-Saharan Africa and proposed that vaccination, past infection history, and effective mitigation strategies might have reduced mortality (*28*–*30*). The age pattern of deaths in lower-income countries has skewed younger than in high-income countries, more so than can be explained by age distribution alone, but the explanation for this fact is not known (*31*–*33*).

Estimating infection fatality rates requires accurate measurement of the number of infections and of deaths in the population due to COVID-19. Widely reported CFRs rely on the number of infections measured by positive tests undertaken largely for clinical purposes. CFRs may dramatically overestimate IFRs, especially in low-income settings where testing capacity is limited (*34*) and therefore tests are reserved for symptomatic patients who are more likely to be infected. IFR estimates almost universally rely upon large-scale seroprevalence samples drawn from the larger population, matched to administrative data on deaths. There have been very few seroprevalence studies (listed above) that can be matched to mortality counts in lower-income settings and none with sufficient sample size to calculate age-specific IFRs with any granularity (*24*). But IFRs that are not age-specific are difficult to compare across contexts, because the age pattern of infection may vary and aggregate IFRs will be larger in places where older people have a larger share of infections.

We use three data sources from India that are uniquely well-suited to calculating age-specific IFRs. We first use population-representative seroprevalence surveys in the city of Mumbai (N∼=7000) and in the state of Karnataka (N∼=1200). By matching results from these surveys to age-specific administrative data on deaths at the time of seroprevalence surveillance, we can calculate IFR without relying on non-representative testing data. Karnataka and Maharashtra (the state of Mumbai) are among the states of India with higher quality epidemiological surveillance and death registration (*35*). The third data source is a survey of COVID-19 prevalence among randomly sampled short-term outmigrants (mostly working-age men) returning home to the state of Bihar, with mortality followup. Because these migrants were randomly sampled and tracked until recovery or death, the death rate among those who tested positive in this group is interpretable as an IFR.

Our objective was to calculate age-specific IFRs in all three locations and to compare them to international estimates, which are based mostly on high-income countries. We further examined heterogeneity of mortality rates across the three sample locations, and by age.

## Methods

In Mumbai and Karnataka, we matched representative seroprevalence surveys to administrative reports of COVID-19 deaths. In Mumbai, seroprevalence surveys were conducted for two weeks in July 2020 on representative samples of three wards, one from each of the city’s three zones, stratified by age, gender and slum/non-slum dwellers (*10*). The sample consisted of 6,904 participants (4,202 from slums and 2,702 from non-slums), tested for IgG antibodies to the SARS-CoV-2 N-protein using the Abbott Diagnostics Architect™ test. After adjusting for test sensitivity, we calculated aggregate seroprevalence for each ward and multiplied by ward population to obtain a count of the number of people infected. We estimated infection counts to non-sampled wards by assuming a constant rate of government under-reporting in wards in the same zone, an approach that was supported by the finding of very similar case-to-seroprevalence ratios in the three wards with seroprevalence data. Age- and sex-specific infection shares were based on the seroprevalence survey. Data on cumulative deaths was collected from daily reports on COVID-19 from the municipal governing body (Brihanmumbai Municipal Corporation, henceforth BMC). We matched seroprevalence-based infection counts to deaths under the assumption that the delay between infection and seroconversion is on average two days shorter than the delay between infection and death (*36, 37*). To implement this, we calculated the IFR as the cumulative number of deaths reported as of two days after the end of seroprevalence testing, divided by the number of infections calculated as above. Sensitivity tests, described in the supplement, were run with alternate assumptions about timing and test sensitivity.

In Karnataka, seroprevalence surveys were conducted from June 15 to August 29, 2020, in representative samples of urban and rural areas in 20 out of Karnakata’s 30 districts, stratified to generalize to five homogenous regions that span all districts of the state (*38*). 1,196 participants were tested with the ELISA test for antibodies to the receptor binding domain (RBD) of the SARS-CoV-2 virus, developed by Translational Health Science and Technology Institute in India. We adjusted for test sensitivity and specificity and used census population counts to aggregate seroprevalence to state-level infection counts, reweighting to match regional age and gender distributions. We collected district-level death data from the Government of Karnataka Department of Health and Family Welfare bulletins. The infection count was matched to the number of deaths reported two days after the last day of seroprevalence surveillance in each region, as above. IFR was calculated as the number of deaths divided by the number of cases identified by seroprevalence surveys. Date matching is described in more detail in the supplement, along with sensitivity tests for results under alternate date assumptions. In particular, because the seroprevalence survey period in Bangalore spanned two months (compared with less than three weeks in the other regions), we show results excluding Bangalore, where deaths may have been overestimated due to the longer survey period.

In Bihar, the state government began COVID-19 testing among returning out-of-state migrants soon after the first positive case was identified in a migrant on March 22. Beginning on May 4, Bihar began to randomly select migrants for testing. Random testing continued until July 21, though for a brief window (May 22–31) only migrants returning on trains from seven major cities were sampled for testing. We isolated the subsample of migrants who were randomly selected for testing, yielding 4,362 individuals with positive tests. Tests were conducted with TrueNat machines manufactured by MolBio Diagnostics in Goa, with positive tests confirmed by real-time PCR kits (*39*). Bihar attempted to track all migrants, obtaining positive tests until they eventually recovered or died. 1,530 infected individuals (35%) could not be tracked. In main estimates, we assumed that their fatality rates were the same as successfully tracked individuals; in the sensitivity analysis, we considered the possibility that all survived. High attrition is common in studies of migrant workers (*40*), with followup in this case complicated by the ongoing crisis. The vast majority of short-term migrants are working-age men; we limited our analytic sample to 3,921 randomly-sampled male migrants, 2,536 for whom outcomes are known.

Representative seroprevalence surveys matched to administrative death data are the primary source of IFR measurement everywhere in the world (*3*–*7*). The surveys in Mumbai and Karnataka thus give us credible and comparable estimates of age-specific fatality rates in those regions. In Bihar, because migrants were randomly sampled, there is no selection on symptomatic or severe cases, and mortality rates among positive cases can be interpreted as IFRs. We found supporting evidence that the sampling was indeed random in that the symptomatic rate among randomly tested migrants was similar to that among those testing positive in the seroprevalence surveys. As we note in the discussion, short-term migrants from Bihar are economically marginalized; their IFRs can be understood as representative of the migrant population, but not necessarily the general population.

We calculated IFRs in 10-year age bins in all locations, as well as for individuals aged 10–49 and 50–89. We used two large-scale meta-analyses (*3, 7*) of age-specific SARS-CoV-2 IFRs as reference groups. Both Levin et al. (*3*) and O’Driscoll et al. (*7*) draw almost exclusively from seroprevalence samples in high-income areas in Europe and the United States. The application of these seroprevalence samples to mortality in LMICs (as in O’Driscoll et al. (*7*)) requires the as-yet untested assumption that multiple epidemiological factors (*e*.*g*., transmission dynamics, infection attack rate) are uniform across HICs and LMICs. Levin et al. (*3*) do not report IFRs separately for men and women; we estimated gender-specific IFRs from Levin et al. by assuming the same relative IFR for men as was reported in O’Driscoll et al. For the larger age bins, we weighted sample populations and meta-analysis age-specific IFR estimates by the Indian national population distribution, to ensure that differences across contexts were driven by differences in age-specific IFRs rather than different distributions of ages within each bin.

We calculated the slope of natural log of the IFR as a function of age, by fitting a linear function to the most granular age-specific IFR data that could be obtained in each location. Additional details on the underlying samples and the methodology can be found in the Supplementary Materials.

## Results

We plotted age-specific IFR for each location on a log scale, to enable comparison at all ages in spite of the exponential increases in mortality at higher ages found in all countries (Figures 1a and 1b). For both men and women, there is substantial variation in IFRs across the three locations in India. In Karnataka, age-specific mortality rates are an order of magnitude lower than those reported in the meta-analyses, especially over age 70. In Mumbai, estimates were close to the lower of the two meta-analyses at younger ages, but then considerably lower in Mumbai at ages above 60. For 60–69-year-old men, for example, we measured an IFR of 0.17% [95% CI: 0.092, 0.240] in Karnataka and 0.62% [95% CI: 0.591, 0.647] in Mumbai (Table 1); the meta-analyses reported male IFR of 1.02% and 1.86% in this age group, respectively.

**Table 1.** Age-specific infection fatality rates from three locations in India

| Age | Karnataka |  | Mumbai |  | Bihar migrants |
| --- | --- | --- | --- | --- | --- |
|  | Male | Female | Male | Female | Male |
| (1) | (2) | (3) | (4) | (5) | (6) |
| 10–19 | 0.000% | 0.001% | 0.004% | 0.001% | 0.000% |
|  | [0.000, 0.000] | [0.000, 0.001] | [0.004, 0.004] | [0.001, 0.002] | [0.000, 0.000] |
| 20–29 | 0.004% | 0.002% | 0.013% | 0.005% | 0.649% |
|  | [0.002, 0.005] | [0.001, 0.002] | [0.012, 0.013] | [0.005, 0.005] | [0.131, 1.166] |
| 30–39 | 0.013% | 0.006% | 0.041% | 0.019% | 1.810% |
|  | [0.009, 0.016] | [0.005, 0.008] | [0.039, 0.042] | [0.018, 0.020] | [0.795, 2.825] |
| 40–49 | 0.027% | 0.012% | 0.112% | 0.058% | 1.529% |
|  | [0.022, 0.032] | [0.010, 0.015] | [0.107, 0.117] | [0.055, 0.061] | [0.199, 2.859] |
| 50–59 | 0.073% | 0.040% | 0.355% | 0.172% | 2.381% |
|  | [0.058, 0.088] | [0.032, 0.048] | [0.340, 0.371] | [0.164, 0.180] | [0.000, 5.043] |
| 60–69 | 0.166% | 0.050% | 0.619% | 0.317% | 4.255% |
|  | [0.092, 0.240] | [0.030, 0.070] | [0.591, 0.647] | [0.302, 0.331] | [0.000, 10.026] |
| 70–89 | 0.163% | 0.106% | 0.837% | 0.511% | 12.500% |
|  | [0.074, 0.252] | [0.051, 0.160] | [0.800, 0.875] | [0.487, 0.534] | [0.000, 35.418] |

Infection fatality rates as percentages. 95% confidence intervals in square brackets.

**Figure 1.**
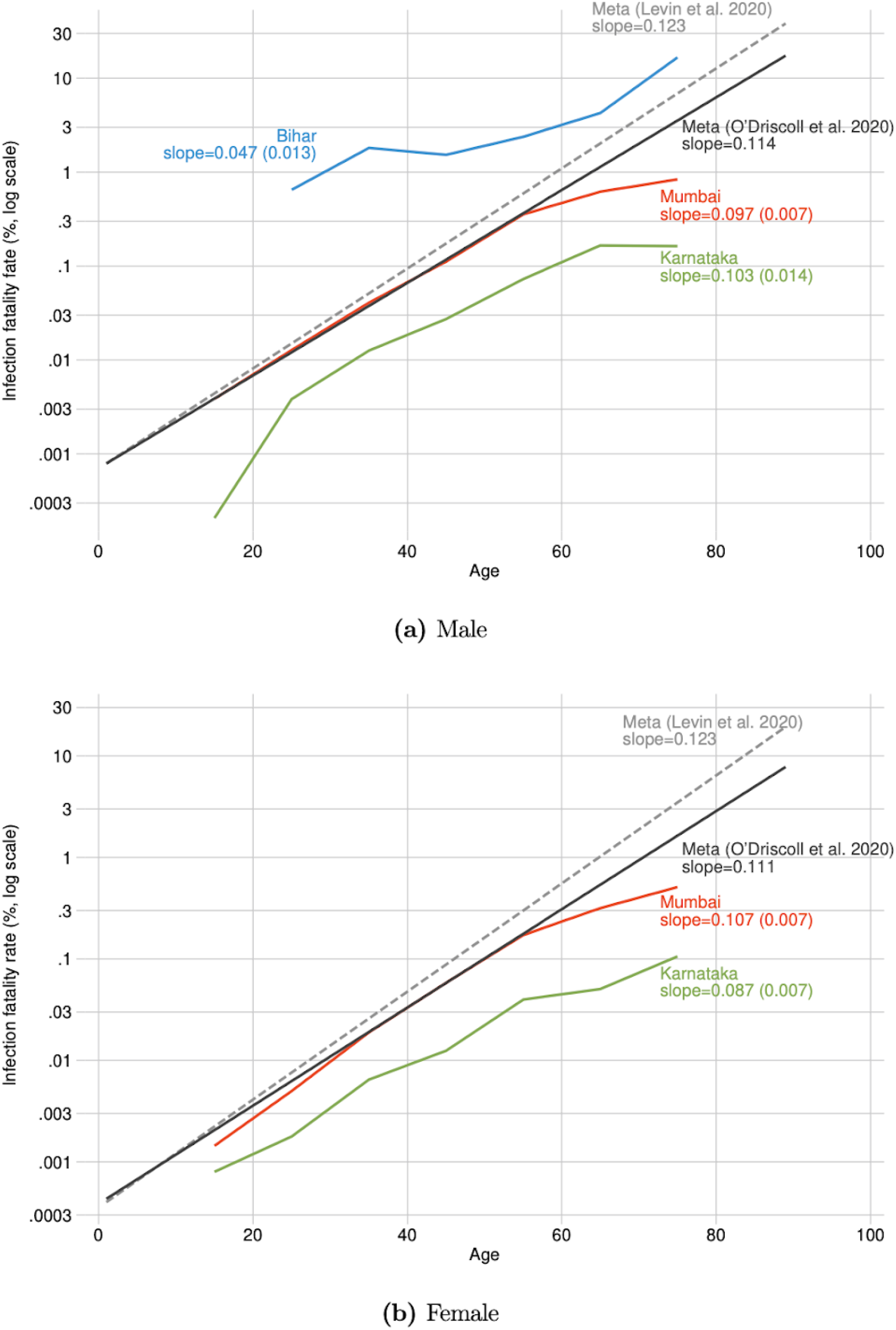
Age-specific infection fatality rate, comparing three locations in India with international estimates

In contrast, mortality among male migrants returning to Bihar is an order of magnitude higher. Mortality among men aged 60–69 was extremely high but measured imprecisely due to the small sample of older men (4.0% [95% CI: 0, 9.4%]). The larger age bins allowed a more precise measure of IFR in Bihar (Table 2). In both the 10–49 and 50–89 age bins, mortality in Bihar was an order of magnitude higher than the other Indian locations and both meta-analyses, after weighting to the Indian age distribution to ensure age-specific comparability. For the 50–89 age group, the estimates were not precise enough to rule out equality between Bihar and the other groups. For the 10–49 age group, we can rule out equality (p < 0.01).

**Table 2.** Age-specific fatality rates in India (ages 10-49 and 50-89)

| Age | Karnataka |  | Mumbai |  | Bihar migrants |
| --- | --- | --- | --- | --- | --- |
|  | Male | Female | Male | Female | Male |
| (1) | (2) | (3) | (4) | (5) | (6) |
| 10–49 | 0.009% | 0.004% | 0.033% | 0.016% | 0.851% |
|  | [0.007, 0.010] | [0.004, 0.005] | [0.032, 0.034] | [0.016, 0.017] | [0.467, 1.235] |
| 50–89 | 0.120% | 0.056% | 0.530% | 0.285% | 5.393% |
|  | [0.090, 0.150] | [0.043, 0.069] | [0.516, 0.544] | [0.277, 0.293] | [0.000, 11.156] |

Infection fatality rates as percentages. 95% confidence intervals in square brackets.

To the extent that an IFR advantage exists in India, it was found among the old more than among the young. In all three regions, the overall increase in IFR with age was considerably less steep than in the reference meta-analyses (Figure 1), particularly at older ages. The meta-analyses suggest that an 80-year-old has about 100x the IFR of a 40-year-old; in Mumbai, the increase in risk factor is 40x and in Bihar it is only 10x. Specifically, male IFR increased on average by 4.7%, 9.7%, and 10.3% with each year of age in Bihar, Mumbai, and Karnataka respectively. We calculated comparable figures in the meta-analyses as 11.4% (*7*) and 12.3% (*3*). The differences between the Indian and the reference groups were similar among women.

The main estimates are replicated in the supplementary materials under a range of different scenarios and assumptions; the ordering of IFRs across regions and with respect to the reference groups is highly robust (Figures 2a-d).

**Figure 2.**
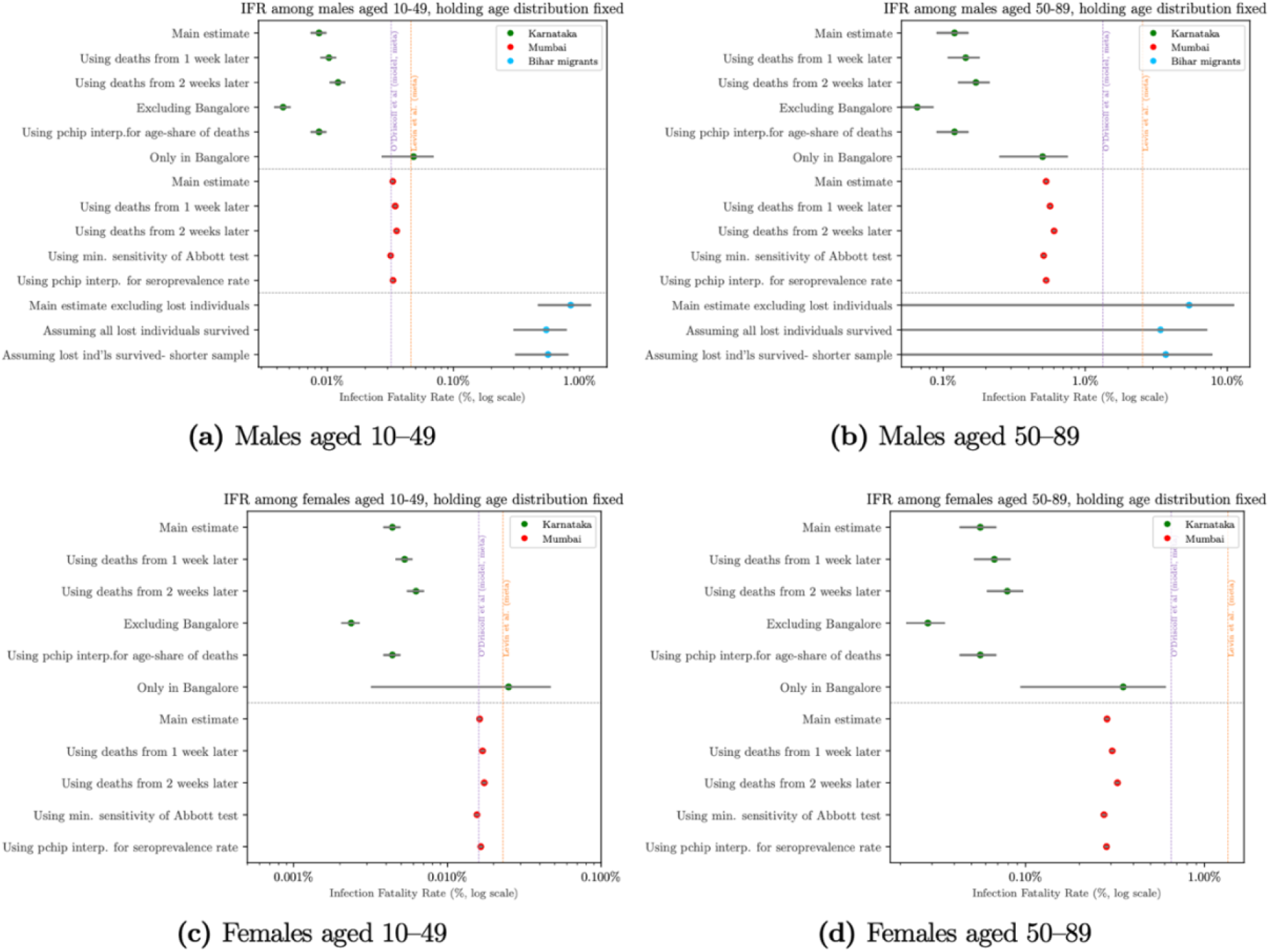
Age-specific infection fatality rates in India: sensitivity tests Markers indicate population-weighted pooled IFRs with India as the reference population. 95% confidence intervals are in grey.

## Discussion

Our results suggest that there is substantial heterogeneity in age-specific IFR from COVID-19 in India. In Mumbai and Karnataka, IFRs were lower than those measured in richer countries, particularly at the ages where most deaths occur. In Bihar, IFR estimates were an order of magnitude higher than the other two locations and the international reference groups.

Migrants to Bihar may have had higher IFRs because they were among the most socioeconomically distressed people in the country. Short-term migrants were on average very poor even before the pandemic, often working in cities in marginal conditions without security of tenure (*40*). The sudden lockdown left them out of work, short of wages, and desperate to return home (*41*). They may even have worse outcomes than slum residents on average (*42*) because they self-selected to leave slums. With transportation options shut down, the return journey was arduous for many (*43*). Migrant workers have worse health than the general population at baseline (*44*); the circumstances at the beginning of the pandemic may have made this group exceptionally vulnerable to adverse health events following viral infection.

Our study also revealed a weaker increase in IFR over age than seen in other countries. It has already been noted that the pattern of mortality in low- and middle-income countries skews younger than would be predicted from age distribution (*33, 45, 46*). Our study suggests that a flatter age profile in mortality could be a major factor driving this difference.

Like all population estimates connected to COVID-19, our estimates are only as good as the underlying data. The largest potential source of bias in this study was the use of official reports of COVID-19 deaths, which are likely to undercount the true number of deaths (*35, 47*).

Misreporting of deaths would have to be substantial, however, to change our conclusions. Focusing on the 50+ age group, in Mumbai, a doubling of COVID-19 deaths would be required to put the IFR in the range of the meta-analyses. It is at least plausible that deaths in Mumbai were undercounted by a factor of 2; from March to July, Mumbai recorded 6,600 excess deaths in addition to the 6,400 COVID-19 deaths used for the estimation in this study (*48*). In Karnataka, COVID-19 deaths would have to be under-reported by a factor of 5 to bring IFR in line with the international estimates. We cannot rule out this kind of death misreporting; however, we calculated IFR using a standard methodology used in many cross-national settings, many of which are also characterized by under-reporting of COVID-19 deaths. Finally, official misreporting of COVID-19 deaths would not bias our IFR estimates in Bihar, due to the mortality followup methodology underlying these estimates. For our Bihar estimates to match the range of meta-analyses, deaths would need to have been *overcounted* by a factor of 2 for ages 50–89 and a factor 10 for ages 10–49.

It is also possible that our estimates could overestimate IFR. Antibodies fade over time, so seroprevalence tests provide an upper bound on the cumulative infection rate (*49, 50*). If the number of infected patients carrying few antibodies at the time of testing is high, then seroprevalence-based IFR estimates will be biased upward.

In Bihar, we cannot rule out that the government did not adhere to its stated random sampling protocol, and may have sampled individuals who were more likely to fall seriously ill and die. However, we found the same symptomatic rate in positive patients in Bihar as was found in the random seroprevalence test samples, suggesting sampling in Bihar was indeed random. Finally, we do not know the base rate of migrant death in the absence of COVID-19, given the hardships faced by returning migrants. If migrant deaths would be high regardless of COVID-19, we may overstate the mortality attributable to COVID-19 in this group.

At the time of writing, these estimates are the best available in a lower-income setting. Improved epidemiological surveillance and accounting of SARS-CoV-2 are critical investments that would improve our ability to understand the fatality risk of the virus in lower-income settings.

## Data Availability

Replication code will be posted in a public repository on Github. The repository will include all data on demographics and COVID-19 deaths by location, seroprevalence aggregates for Mumbai and Karnataka, and mortality rates by age and gender for migrants from Bihar. We do not have permission to share seroprevalence microdata. Replication code will be provided to reconstruct all results in the paper from these data.

## Acknowledgments

We thank Sabareesh Ramachandran for pointing us toward case-level death data in Karnataka.

## Funding

This paper was partially funded by Emergent Ventures grant #466.

## Author contributions

All authors participated in idea generation and development, empirical strategy design, and manuscript development. Malani and Tandel provided data on seroprevalence and mortality, and contextual knowledge regarding government sampling schemes and mortality registration. Cai and Novosad conducted the data analysis.

## Competing interests

Authors declare no competing interests.

## Supplementary Materials

Materials and Methods

Figures S1-S10

Tables S1-S2

References (51-57 in list above)

## Materials and Methods

### Bihar

#### Data

We made use of data on all positive cases in the state of Bihar found during random testing of incoming migrants during an early phase of the pandemic. The data was provided by the Health Department of the Government of Bihar. The data contained a sample of 4,954 active infections and their outcomes, reported between March 22 (the date on which the first positive case in Bihar was detected) and July 21, 2020. The vast majority of the sample (over 99%) consisted of migrants travelling from within India into Bihar, most on designated trains. Migrants were more likely to be sampled if they presented symptoms between March 22 and May 3. State policy beginning May 4 during the sample collection period mandated that travellers from within or outside India (mainly migrant workers returning home due to travel restrictions) be randomly sampled and tested for COVID-19 infection from March 20 to May 22, and after May 31. Between May 22–31, only migrants from seven high-infection cities (National Capital Region, Mumbai, Ahmedabad, Pune, Surat, Kolkata, and Bangalore) in India were randomly sampled. We isolated the subsample of migrants who were randomly selected for testing, yielding 4,362 cases.

During the sample period, migrants were tested with TrueNat machines manufactured by MolBio Diagnostics in Goa (India), and positive tests were confirmed with real-time reverse polymerase chain reaction (RT-PCR) kits *(53)*. Importantly, all infected migrants were tracked by the monitoring team, to determine whether they eventually recovered or died. Among randomly sampled male migrants, 1,385 infected individuals (35.3%), whom we call “lost”, could not be tracked and thus their final outcome is uncertain. The high level of attrition is common in studies of migrant workers, whose frequent movement complicates administrative registration and tracking, particularly during a crisis (40). We considered several approaches to adjusting for attrition, described below. The migrant sample, reflecting typical labor migration patterns in India, was overwhelmingly male (90%). Thus we limited our final analytical sample to 3,921 randomly sampled male migrants, for 2,536 of whom outcomes (recovery or death) are known.

#### Estimating infection fatality rate

Because everyone in the sample had tested positive for SARS-CoV-2, IFRs were estimated as the share of deaths among non-lost individuals in each age group. To account for potential biases due to attrition and delays between infection and recovery/death/reporting, we estimated IFRs using three separate methods, and report estimates from all three.

In age group *a*, denote the number of lost cases as *n*_*a,lost*_, the number of recovered cases as *n*_*a,recovered*_, and the number of cases ending in death as *n*_*a,died*_.

Method 1 (main estimation): In our main estimation, we assumed that lost cases had the same IFR as successfully tracked cases, within each age group. This assumption was implemented by excluding lost individuals from the IFR calculation. Method 1 provided a midline IFR estimate:

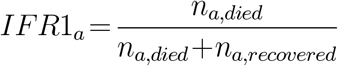

Method 2:In this estimation, we assumed that all lost cases eventually recovered. Thus Method 2 provided a lower bound IFR estimate:

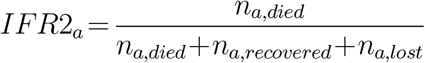

Method 3: The share of cases with successful followup declined in late July as the volume of migrants increased. To account for potential right-censoring of reported outcome rate due to delays between report of initial infection and report of recovery/death, in the third method, we dropped all cases reported within two weeks of the last report date (July 21st):

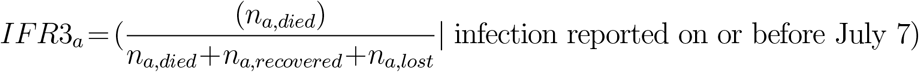

Standard errors were estimated with the normal approximation for a proportion from multiple draws from a binomial distribution.

### Mumbai *Data*

Data on seroprevalence were obtained from a representative, stratified, random sample of slum and non-slum populations in three of twenty-four wards of Mumbai (see Malani et al. *(10)* for full survey design). Sample collection lasted two weeks and ended on July 14th in slums and July 19th in non-slums. The three wards were selected to represent the city’s three broad zones (city, eastern suburbs, western suburbs); choice of sampled ward within each zone was by convenience. The sample consists of 6,904 participants (4,202 from slums and 2,702 from non-slums), who were tested for IgG antibodies to the SARS-CoV-2 N-protein using the Abbott Diagnostics Architect ™ N-protein based test. The samples were stratified by four age groups, gender, ward, and slum/non-slum residence.

Data on reported infections and deaths by ward and age distribution of deaths were provided in reports released by the municipal governing body (Brihanmumbai Municipal Corporation, hereafter BMC). Data on ward population in slums and non-slums came from the 2011 Population Census. Data on shares of population by age and gender in each ward-slum came from the 2012 Socio-Economic and Caste Census.

### Estimating IFR

#### Estimating number of infections

The seroprevalence survey reported seropositivity in four age groups (12–24, 25–39, 40–60, 61+), called “coarse bins”. To generate infection counts that could be compared with city death statistics (which are reported in 10-year age bins), seropositivity by 10-year age bin was interpolated by fitting a non-linear function over seropositivity in the coarse bins. For the main estimation, we interpolated seropositivity in 10-year bins, using the inverse distance-weighted mean of non-missing values (using the Stata package mipolate), weighting with the squared inverse of distance. In each coarse bin, the median age of residents in Mumbai City was used as the non-missing value for age. Figure S1 shows the observed and interpolated values. As a sensitivity analysis, we report IFR estimates using a piecewise cubic Hermite (“pchip”) interpolation for seropositivity (see Figure S2 for visual). Interpolation predicted seroprevalence for the midoint of each 10-year age bin, separately by gender, ward, and slum status.

The estimated sensitivity of the chemiluminiscence immunoassay ranges from 90% (95% CI: 74.4%–96.5%) (56) to 96.9% (95% CI: 89.5%–99.5%) *(52)* while specificity in those studies was 100% (95% CI: 95.4%–100%) and 99.90%, respectively. We estimated seroprevalence from seropositivity using the Rogan-Gladen correction (*(54)* to account for imperfect accuracy of tests. In the main results, we used the midpoint of mean sensitivity estimates (93.45%) and the midpoint of corresponding specificities (99.5%). As a sensitivity analysis, we replicated results with an upper bound for seroprevalence based on the Abbott test’s lower bound of sensitivity (90.0%) and upper bound of specificity (100%) (52) (Figure S7).

Denote the estimated number of infections in age bin *a*, gender *g* in sampled ward *s* as:

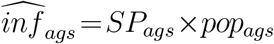

where *SP*_*ag,s*_ is the estimated seroprevalence rate, and *pop*_*ag,s*_ is population.

#### Estimating the number of infections in non-sampled wards

BMC death data reported the ward of death, but not the ward of residence. Discussion with government officials and review of the data indicated that the ward of death was not a reliable indicator of ward of residence. This implied that calculating IFR by dividing the number of ward-level deaths by the number of ward-level infections would overestimate deaths in wards with large hospitals and underestimate them elsewhere. Instead, we used the seroprevalence surveys to generate estimates of city-wide infection counts.

To estimate true number of infections in non-sampled wards, we drew on administrative ward-level infection counts (which were universally available from city reports), and assumed that they were proportional to actual infections at similar rates in different wards of the city. Effectively, this amounts to assuming that the BMC underestimated the true population infection count at the same rate in sampled and non-sampled wards within the same zone. This assumption is supported by Table S1, which shows that in the three wards where we obtained seroprevalence data, case multipliers were very similar.

Thus, in each zone *z*, we calculated a case multiplier based on sampled ward *s*:

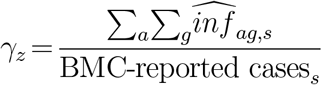

The multiplier indicates the under-reporting rate in each zone *z*. The numerator of the expression is calculated from the seroprevalence surveys as above, and the denominator is taken from the BMC reports. BMC-reported cases were measured as of July 19, the last day of seroprevalence sample collection. We then multiplied the BMC’s reported number of positive cases in non-sampled ward *n* in zone *z* by *γ*_*z*_. That is,

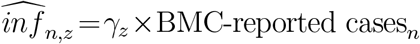

The benefit of this approach is that it allows pandemic intensity to vary across wards, a realistic assumption given significant ward-level variation in reported cases per capita and in the number of containment zones.

This approach also implicitly assumes that the BMC under-reports cases in slums and non-slums at the same rate, *i*.*e*. a ward’s case multiplier does not depend on share of population living in slums. This assumption is also supported by the consistent multipliers reported in Supplement Table S1, across three wards with different slum shares.

#### Estimating the number of infections in each age-gender group in non-sampled wards

We did not observe the age and gender distribution of infections outside of the sampled wards, so it was necessary to assume that non-sampled wards had the same age and gender distribution of infections of sample wards. This was supported by similar age and gender distributions of infections in the three wards with seroprevalence surveys. Figure S3 shows the calculated age and gender distribution of infections; note that the distribution of infections measured with seroprevalence skews younger than the number of reported positive cases, which we presume omits many infected but asymptomatic young people. This approach would cause error if the age distribution varied substantially across wards, but it is overall quite similar; even the median age gap between slums and non-slums was less than one year.

The number of infections in non-sampled ward *s* for gender *g* in age *a* was thus calculated as:

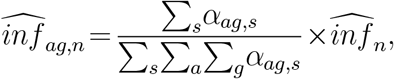

where *α*_*ag,s*_ is the age-gender group’s share of total cases in sampled ward *s*.

#### Estimating the number of deaths

To map seroprevalence numbers to death numbers, the time between infection and death and the time between infection and seroprevalence are needed. The literature suggests a distribution of delay between symptom onset and death *(36)* that is wider than that between onset and seroconversion *(37)* Linton et al. estimated a median time delay of 13 days (17 days with right truncation) between illness onset to death. Stringhini et al. estimated a mean delay of 11.2 days between symptom onset and seroconversion. Based on these estimates, we assumed that the delay between infection and death is on average two days longer than the delay between infection and seroconversion. In the main results, the number of deaths was therefore measured as the cumulative deaths reported in each Mumbai ward as of July 21. This is likely to slightly overstate the IFR, since some deaths may have been associated with individuals who contracted the virus after testing negative in the seroprevalence surveys. However, this upward bias is partially balanced out by the fact that the time between seroconversion and death is not uniform and is likely to be longer than 2 days for a non-trivial share of cases.

Rather than model non-uniform delays between infection and death, we bounded our IFR estimates from above by choosing more conservative death dates. In sensitivity analyses reported below (Figure S6), we replicated IFR estimates using deaths from one week (July 28) and two weeks (August 4) after the end of seroprevalence surveying, both of which plausibly overestimated the number of deaths related to the seroprevalence surveys, given the context of steadily increasing case counts in Mumbai from June to August.

The assumption that deaths measured 1 and 2 weeks later will lead to upward biased IFRs is further strengthened by recent evidence from roughly 125,000 cases in two other Indian states, which found that delays between case report and death were significantly shorter than delays found in China *(45)* and the United States *(57)*.

We used the age distribution of deaths as reported by the BMC up to the date used for measuring deaths, and the gender distribution (65% male, 35% female) up to August 3 *(51)* (the gender distribution of deaths was not included in earlier reports). This yields the estimated number of city-wide deaths by age-gender group, *d*_*ag*_.

### Estimating city-wide IFR by age in Mumbai

Denote the final city-wide IFR in Mumbai, in age bin *a* for gender *g*, as *IFR*_*ag*_:

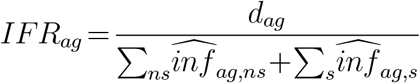

Standard errors of IFRs were calculated reflecting propagation of the design-based standard errors of the age- and gender-specific seroprevalence estimates with a normal approximation.

### Karnataka

#### Data

Data on seroprevalence were obtained from the Karnataka Seroprevalence Survey (hereafter KSS) a state-wide representative sample of urban and rural areas in 20 out of 30 districts in Karnataka, representing 5 broader regions (see Mohanan et al. *(38)* for a detailed survey description). The sample was collected from June 15 to August 29, 2020. Collection times within individual regions were significantly shorter. The study sample was drawn from an existing representative sample of a panel survey—the Consumer Pyramids Household Survey (CPHS)—collected by the Center for Monitoring Indian Economy (CMIE). Our analytical subsample consists of 1,196 tests for IgG antibodies to the receptor binding domain (RBD) of the SARS-CoV-2 virus using an ELISA test developed by Translational Health Science and Technology Institute, India. The sample was not stratified by age and gender, an issue addressed below.

Data on confirmed COVID-19 deaths by district were drawn from Government of Karnataka Department of Health and Family Welfare bulletins, which are released several times per week. Data on the age distribution of total COVID-19 deaths were given by public reports from the state COVID-19 task force. Data on the gender distribution of deaths by age group were obtained from an individual-level dataset of confirmed COVID-19 deaths which was updated through July. The case-level death data were parsed from covid19india.org. Age- and gender-disaggregated population for districts and regions was drawn from the 2012 Socio-Economic and Caste Census (SECC).

### Estimating IFR

#### Estimating the number of infections

The KSS dataset was designed to be representative of 5 broader regions in Karnataka. We therefore can take the ELISA positive test rate as an unbiased measure of the region-level positivity rate. We pooled the data across regions to obtain a statewide test positivity rate in each age and gender group, weighting by region population in each age/gender group.

We then corrected for the sensitivity (84.7%) and specificity (100%) of the ELISA imunoassay *(55)* using the Rogan-Gladen correction *(54)*. This yielded the estimated seroprevalence by age-gender group *SP*_*ag*_, which is multiplied by population *pop*_*ag*_ in each age/gender bin to generate an estimated number of infections 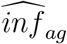, as was done in Mumbai.

### Estimating the number of deaths

The seroprevalence samples were collected at different times in different regions, with the survey period spanning roughly two months (Table S2). To estimate an IFR, we need to match the timing of deaths to the timing of seroprevalence surveying in each region.

#### Choice of dates for measuring deaths

As in Mumbai, we worked from an assumption that the average time difference between seroconversion and death was two days, while testing sensitivity to alternate assumptions (Figure S8). We therefore matched the estimated number of infections calculated in each region to the number of deaths recorded in administrative data two days after the last date of seroprevalence surveying. As in Mumbai, if the two-day delay between seroconversion and death was uniform, this approach would overestimate the IFR, because it counts the deaths of some people who may have been infected *after* recording negative seroprevalence tests.

In all regions except Bangalore, seroprevalence surveying was conducted over a three week period or less, making it straightforward to match test data to death data. In Bangalore, surveying was begun in mid-June but was interrupted by a lockdown. Survey teams returned to finish sampling in the last week of August. Matching Bangalore deaths to the last date of seroprevalence surveying is therefore likely to overestimate the IFR, because a number of those deaths may have been associated with individuals contracting SARS-CoV-2 after testing negative. It was not possible to disaggregate the early and late surveys because death reporting was at the district level, and the early and late survey groups were not representative in and of themselves. To adjust for increased uncertainty regarding the number of infections in Bangalore, we therefore report a sensitivity analysis for all of Karnataka excluding Bangalore (Figure S9).

On some days, official deaths were not reported; in those cases, we used deaths from the following day.^1^

#### Estimating the number of deaths in each demographic group

The Karnataka state government released total death counts on a daily basis, but only intermittently published the age distribution of state-wide deaths. To attribute daily deaths to age and gender groups, we used the age distribution of deaths from the nearest date that was available. The largest period between the date used for deaths and the date used for age-shares was 13 days.

Government reports provided age shares of deaths in 10-year bins in the form (e.g.) 51-60, while the seroprevalence surveys provided age bins in the form (e.g.) 50-59. To harmonize the age groups, we use the medians of the provided bins (e.g. median of 51-60 is 55.5) to interpolate death data to match the age bins in the seroprevalence data, using an inverse distance weighted average method via the mipolate Stata package. Because the target age bins were very close to the available age bins, the risk of error here is small. The fit of the interpolation is displayed in Figure S4). As a sensitivity test, we replicated IFRs using piecewise cubic Hermite interpolation (see Figure S5 for the interpolation and Figure S10 for IFR estimates under the cubic interpolation. For more details, see the discussion on interpolation in Mumbai.

In the absence of death data disaggregated by age and gender on most dates, we assumed that, within age group, the gender distribution of deaths was uniform across regions and equal to the state-wide gender distribution of deaths reported between April and July. This assumption is supported by the finding that IFRs among men were approximately double those among women, consistent with reports from other countries.

Standard errors of IFRs reflect propagation of design-based standard errors of the age- and gender-specific seroprevalence estimates with a normal approximation.

**Figure S1.**
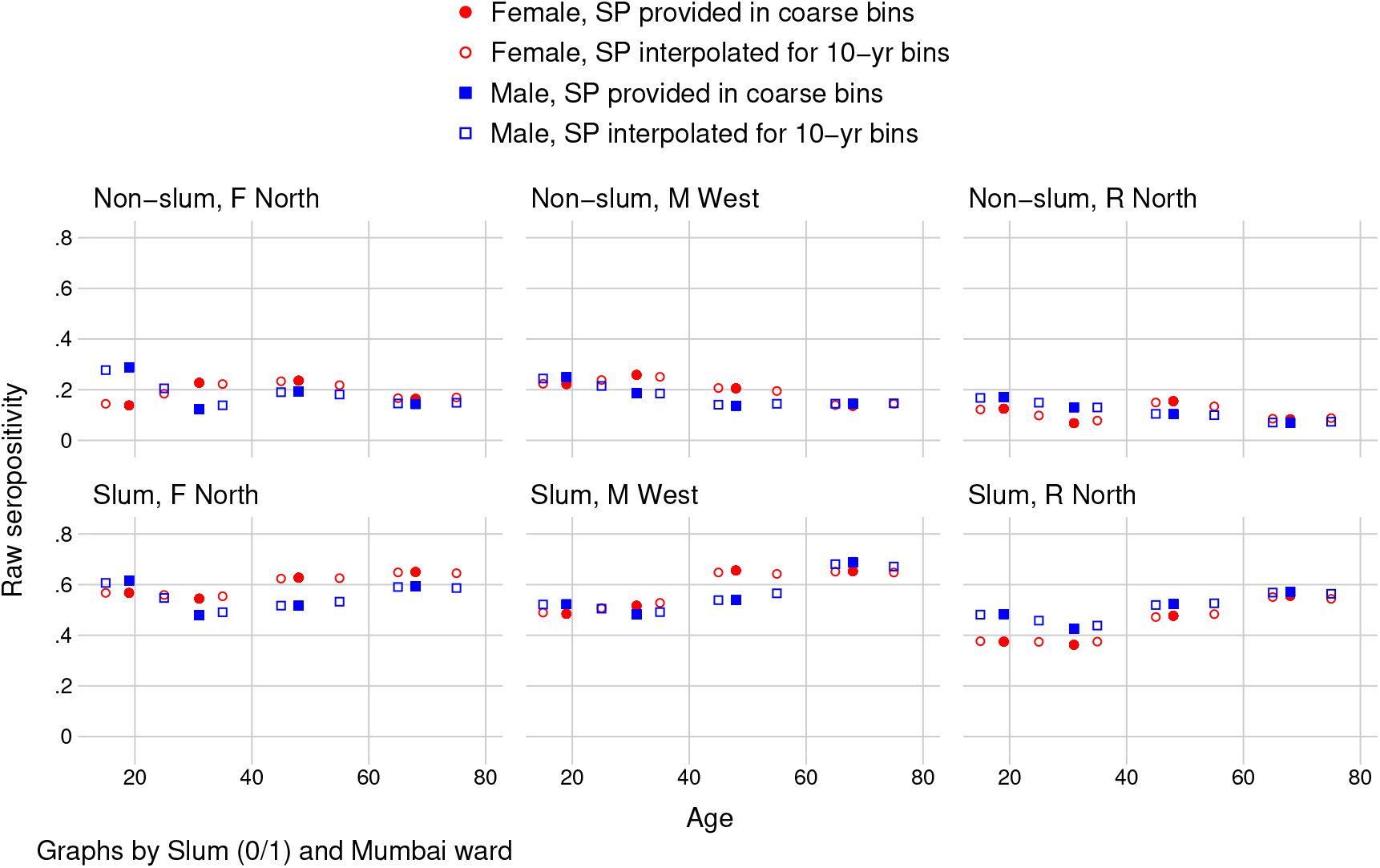
Mumbai: Interpolation of ward-slum seropositivity using inverse distance weighted average

Seropositivity data was provided in four coarse age bins 12-24, 25-39, 40-60, 61+. The figure shows the result of interpolating seropositivity in 10-year age groups from coarse groups using an inverse distance weighted average of known values. All values were interpolated using the median of the age group. Solid markers indicate seropositivity in coarse bins, and hollow markers indicate the interpolated values, to match the age groups in the BMC’s deaths data.

**Figure S2.**
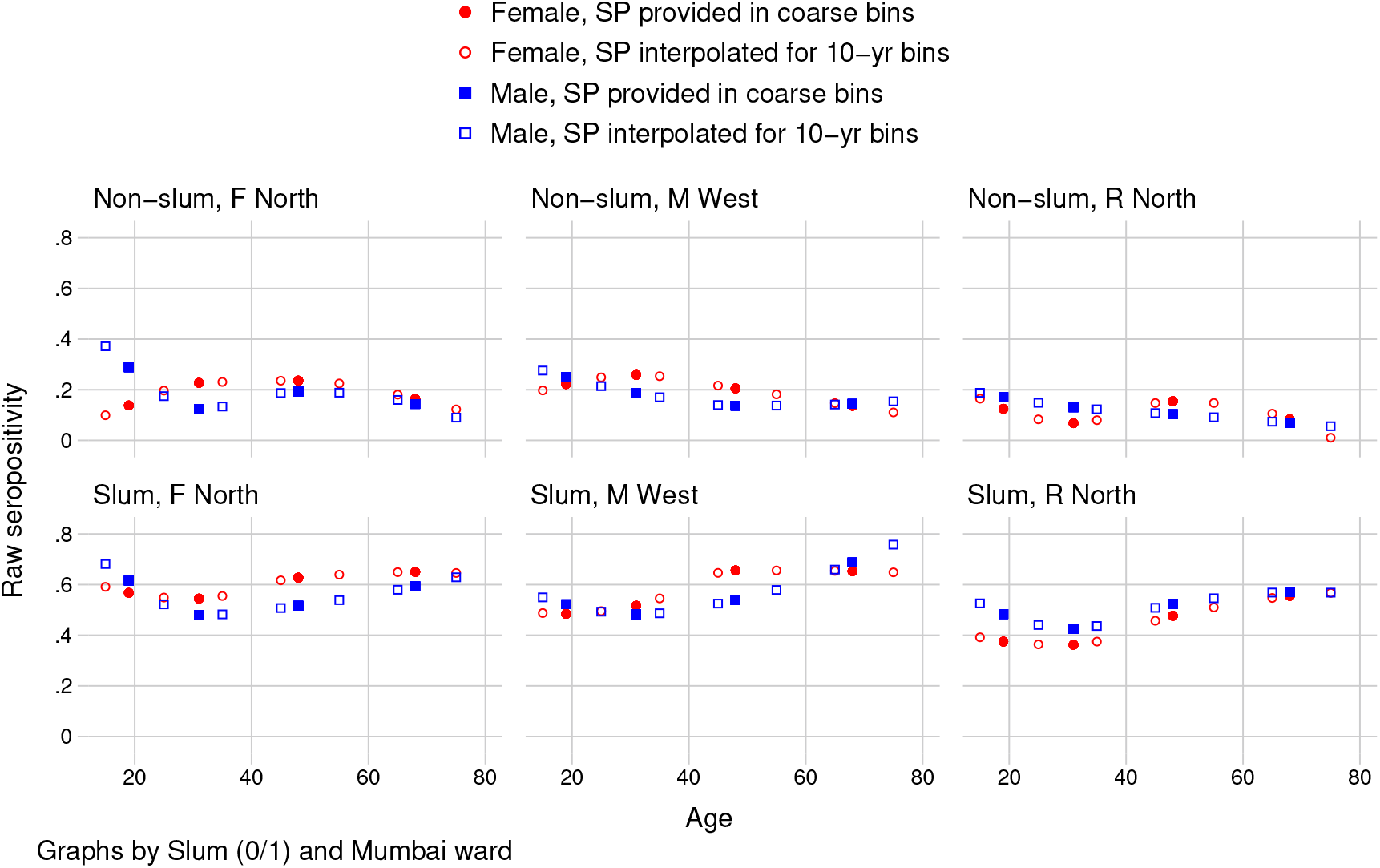
Mumbai: Interpolation of ward-slum seropositivity using piecewise cubic Hermite function

The figure is the same as Fig. S1; the sole difference is that it fits a piecewise cubic Hermite function for interpolation.

**Figure S3.**
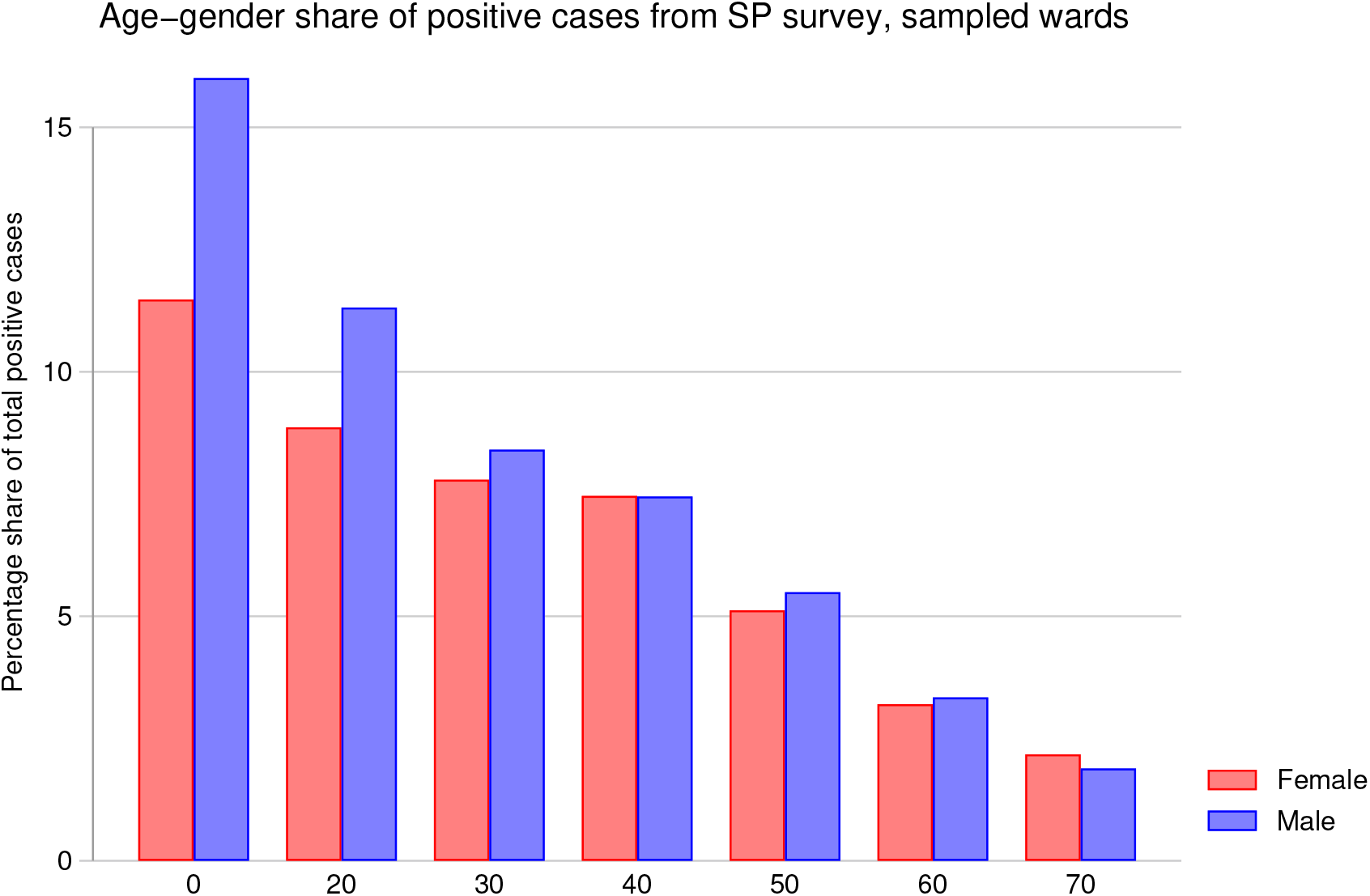
Age-gender cohorts’ share of positive cases from sampled wards in Mumbai seroprevalence survey

“Total positive cases” refers to the estimated number of total infections in Mumbai, multiplying age- and gender-specific seroprevalence rate by group population, summed across age-gender groups and wards. The age- and gender-share of total cases refers to estimated number of infections in age-gender group *ag*, divided by estimated total infections. Age bins are 0-19, 20-29, …60-69, and 70+.

**Figure S4.**
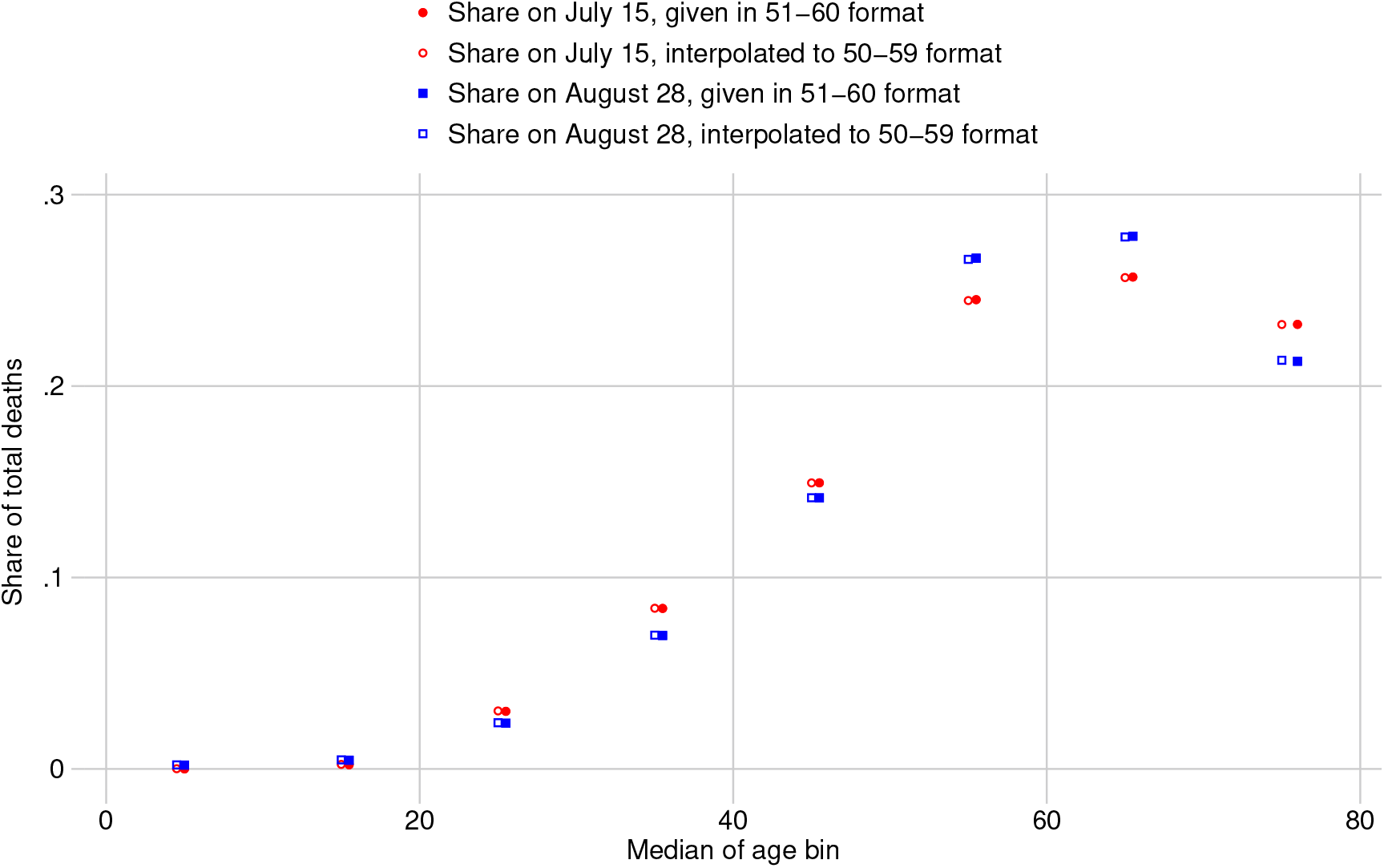
KA: Interpolation of agebin share of deaths using inverse distance weighted average

Data on age-shares of state-wide deaths were provided by the Karnataka Government in age groups of the form 0–10, 11–20, 21–30, etc. The figure shows the result of interpolating age-share of deaths in age groups of the form 0–9, 10–19, 20–29, etc, to match age groups of the seroprevalence data. The interpolated values were estimated using an inverse distance weighted average of the given values. Because we use the age-shares of deaths closest to the date of measured seroprevalence in each region, two dates near the beginning and end of sample collection are provided. Solid markers indicate given values, while hollow markers indicate interpolated values.

**Figure S5.**
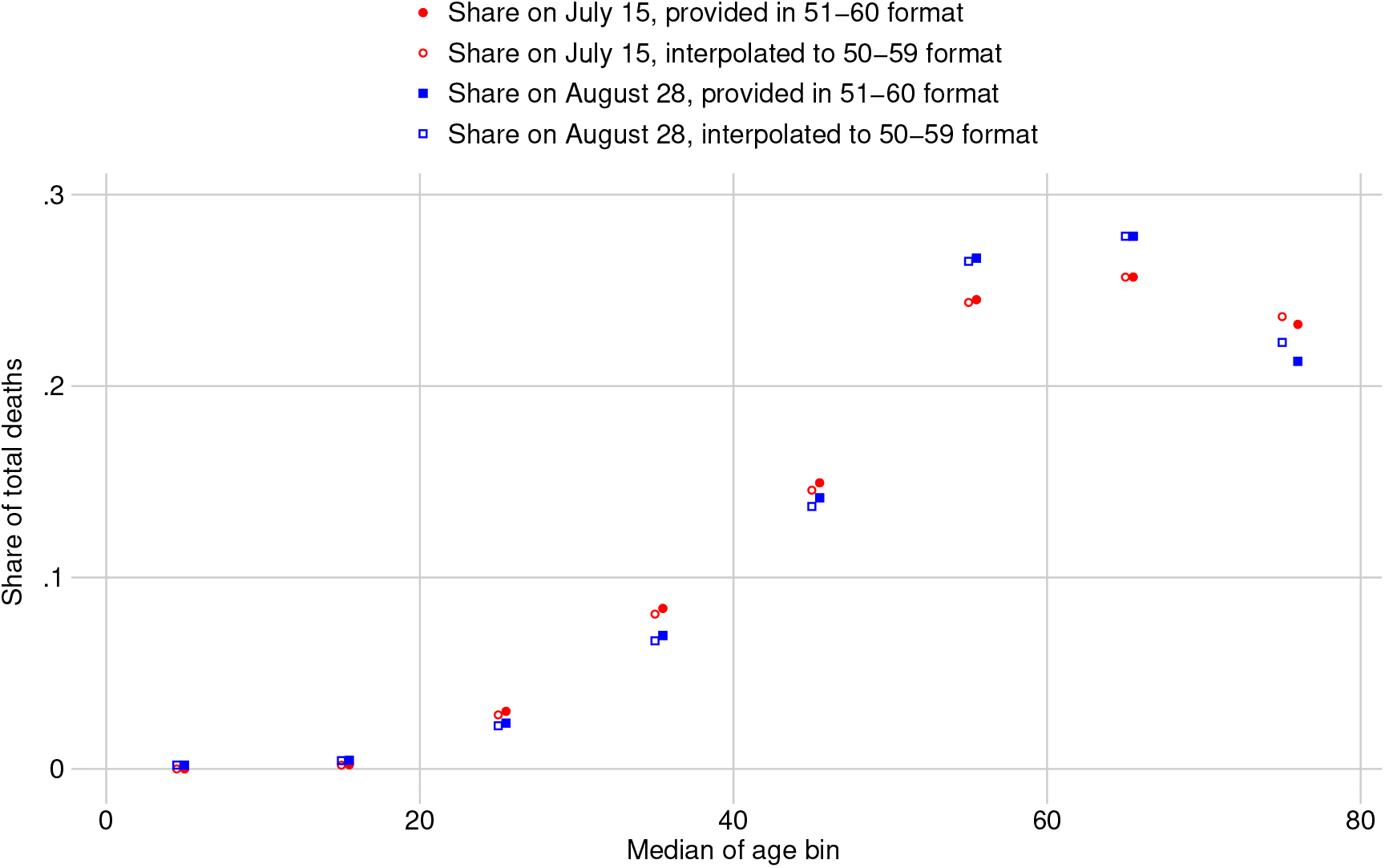
KA: Interpolation of agebin share of deaths using piecewise cubic Hermite function

The figure is the same as Fig. S4; the sole difference is that it fits a piecewise cubic Hermite function for interpolation.

**Figure S6.**
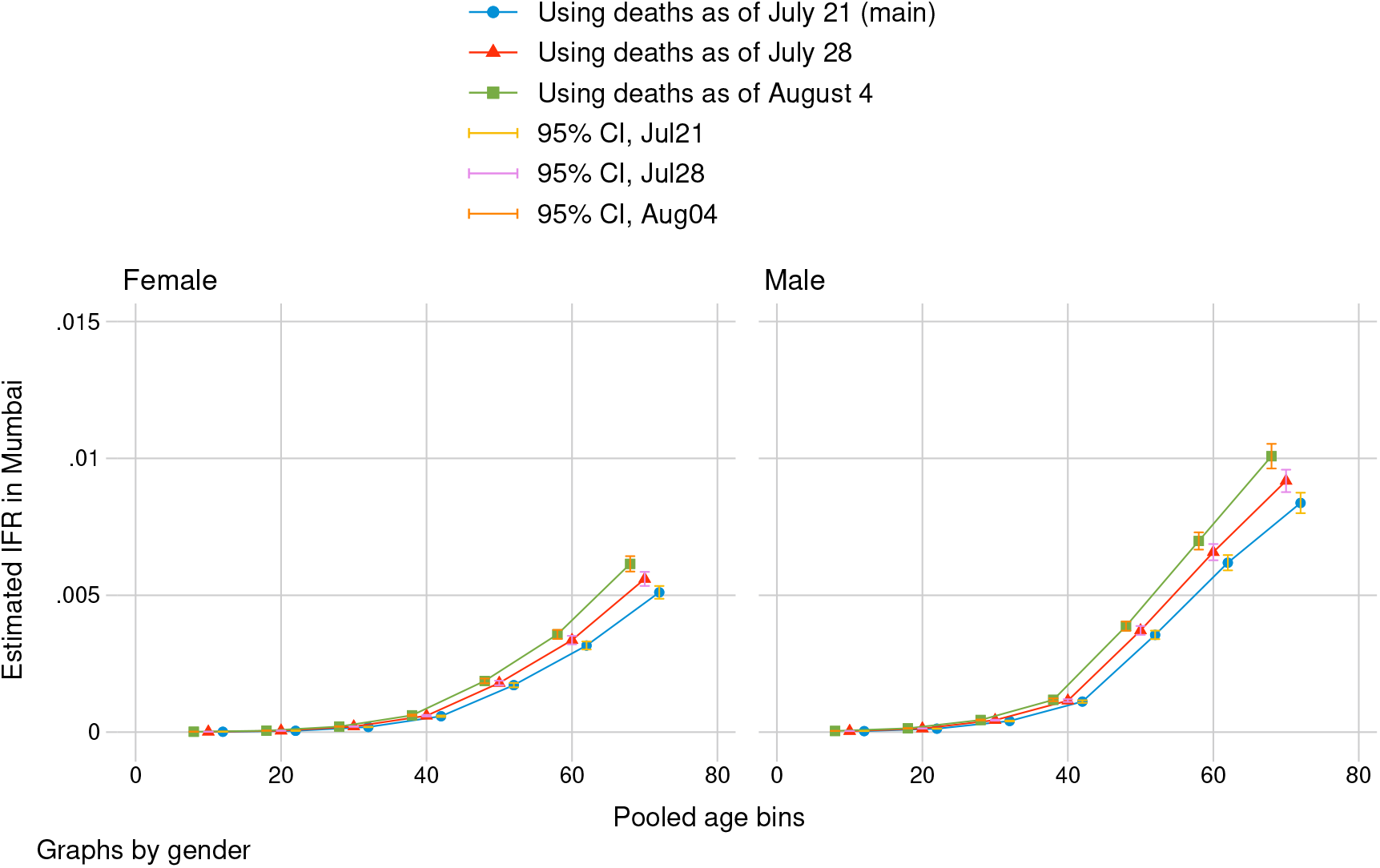
Mumbai: sensitivity analysis using number of COVID deaths from 1 and 2 weeks after date of deaths in main estimation

“Date of death” refers to the day on which we measured cumulative deaths as reported by the city government (BMC). The main date specification measured deaths two days after the end of seroprevalence sample collection. Graphs by gender with 95% confidence intervals. Standard errors reflect propagation of error from design-based uncertainty of seroprevalence estimates. IFRs are calculated in age bins 0-19, 20-29, … 60-69, and 70+.

**Figure S7.**
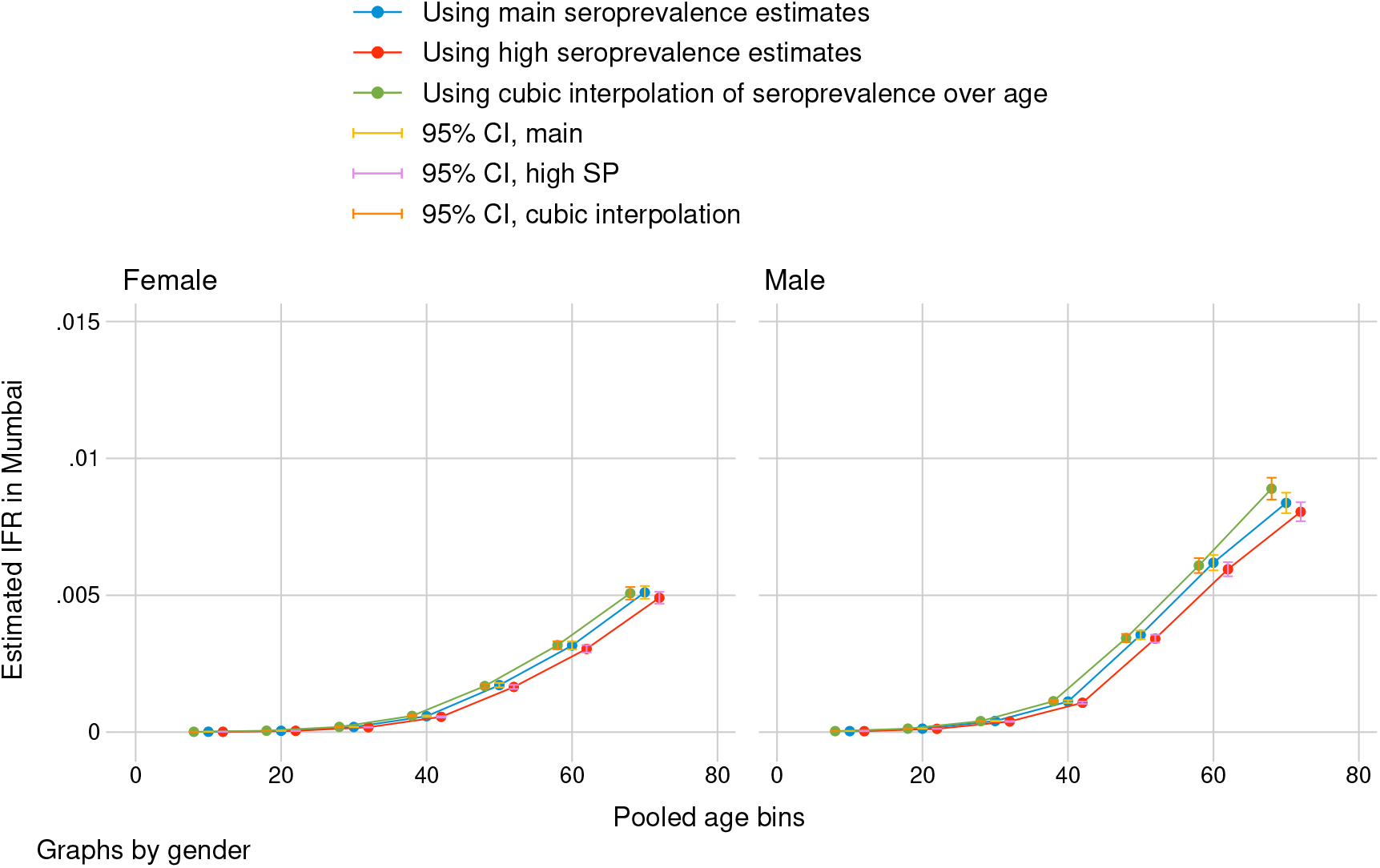
Mumbai: sensitivity analysis, using alternative estimate of seroprevalence and different interpolation method

“Main seroprevalence estimates” use midpoint sensitivity estimate of the Abbott antibody test to calculate seroprevalence from seropositivity in sampled wards, then interpolates seroprevalence to finer age bins with inverse distance weighting (IDW). “High seroprevalence estimates” use minimum sensitivity of the Abbott test to calculate seroprevalence from seropositivity and IDW interpolation. The final sensitivity analysis uses midpoint sensitivity, but piecewise cubic Hermite interpolation to estimate seroprevalence in finer bins. Graphs by gender with 95% confidence intervals. Standard errors reflect propagation of error from design-based uncertainty of seroprevalence estimates. IFRs are calculated in age bins 0-19, 20-29, … 60-69, and 70+.

**Figure S8.**
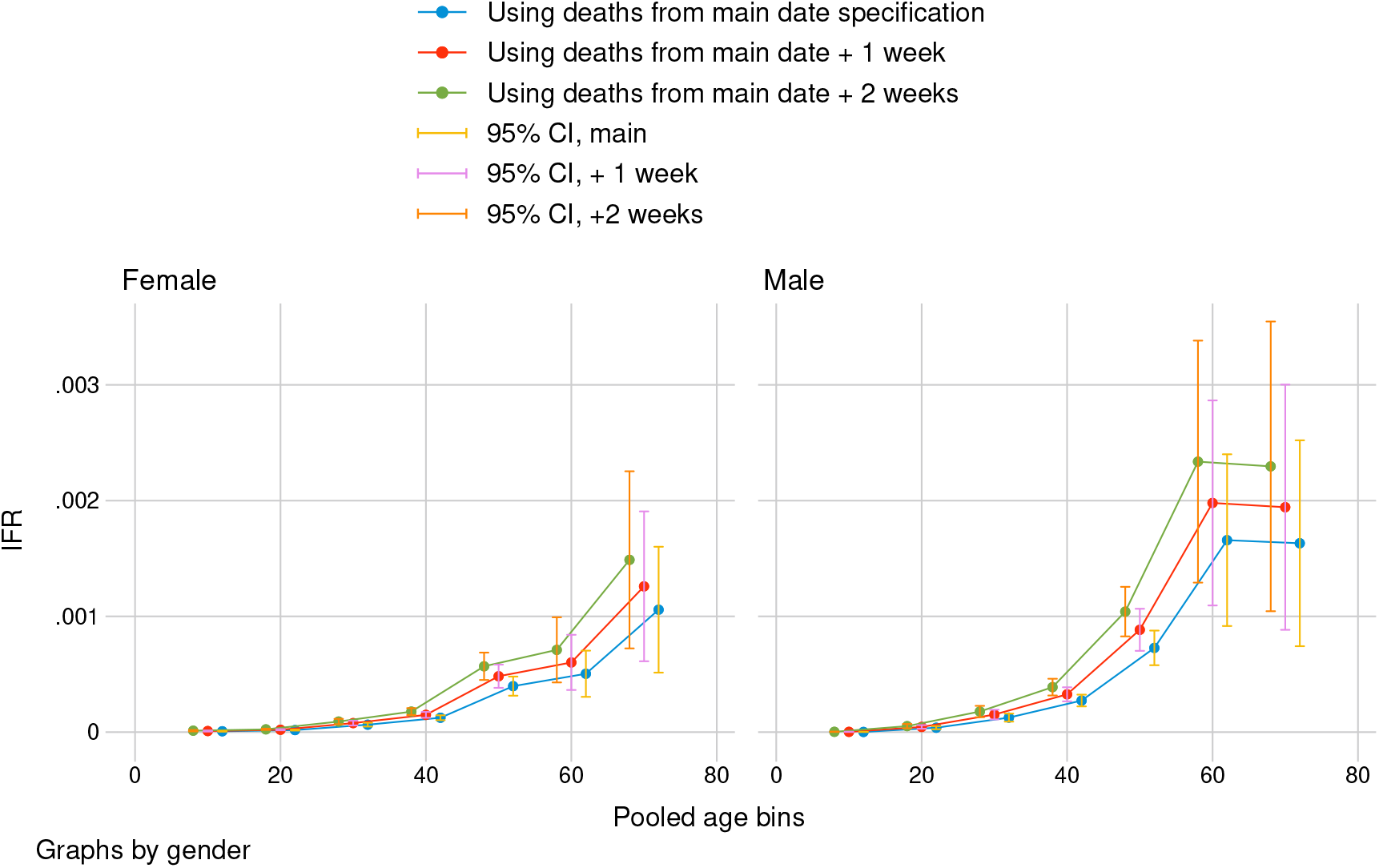
Karnataka: sensitivity analysis using number of COVID deaths from 1 and 2 weeks after date of deaths in main estimation

“Date of death” refers to the date on which we measured cumulative COVID-19 deaths. Main date of specification was determined separately for each sampled region: two days after the median date of sample collection if sampling duration exceeded 21 days, and two days after the last date of collection otherwise. Graphs by gender with 95% confidence intervals. Standard errors reflect propagation of error from design-based uncertainty of seroprevalence estimates. IFRs are calculated in age bins 0-9, … 60-69, and 70+.

**Figure S9.**
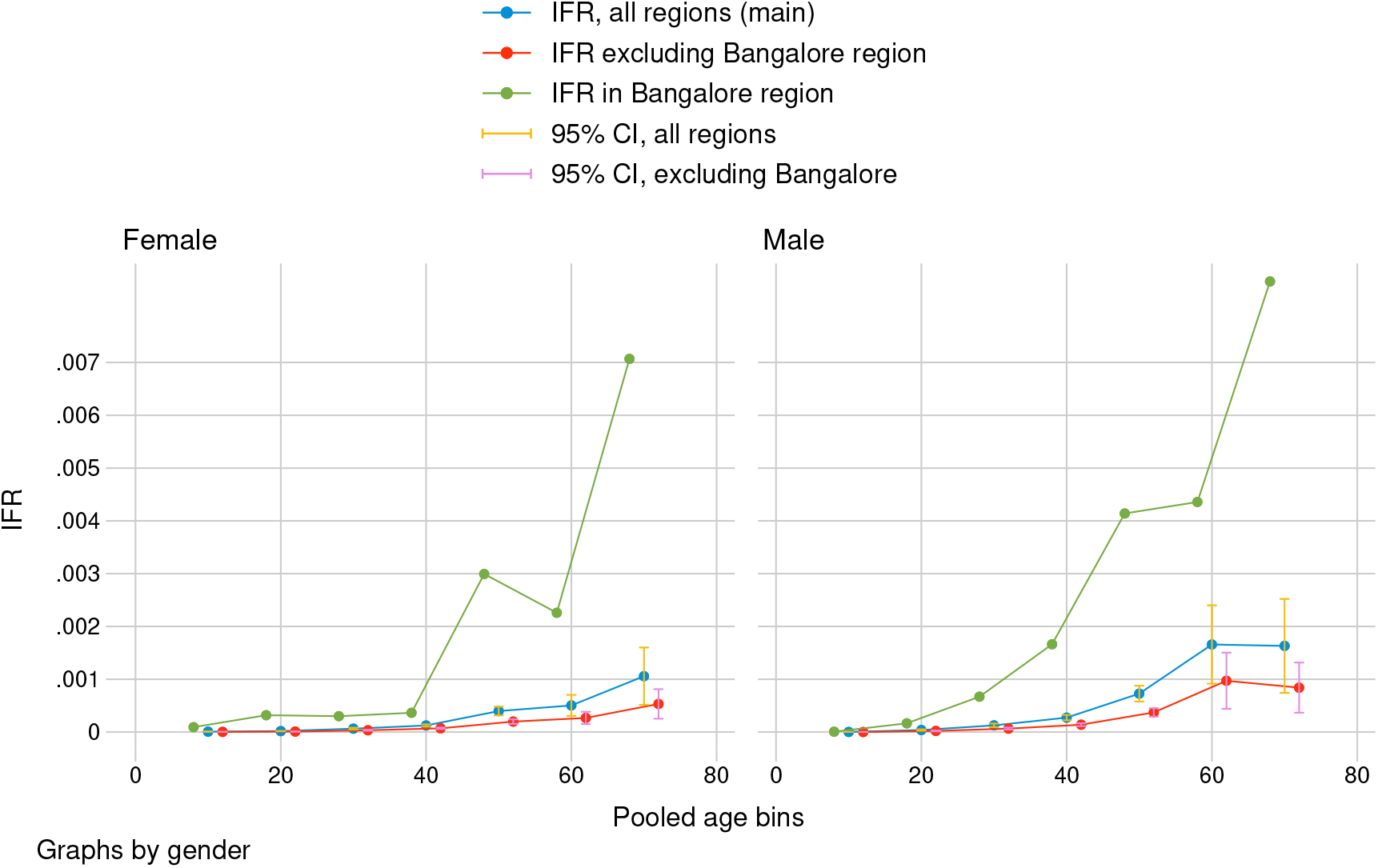
Karnataka: sensitivity analysis isolating Bangalore from other sampled regions

IFRs in main specification are calculated by pooling seroprevalence and death estimates from all five sampled regions of Karnataka. IFRs excluding Bangalore pool from the four remaining regions. Graphs by gender with 95% confidence intervals. Standard errors reflect propagation of error from design-based uncertainty of seroprevalence estimates. Confidence intervals are not reported for Bangalore due to small sample size, and age-specific estimated IFRs in Bangalore should not be interpreted as conclusive. IFRs are calculated in age bins 0-9, … 60-69, and 70+.

**Figure S10.**
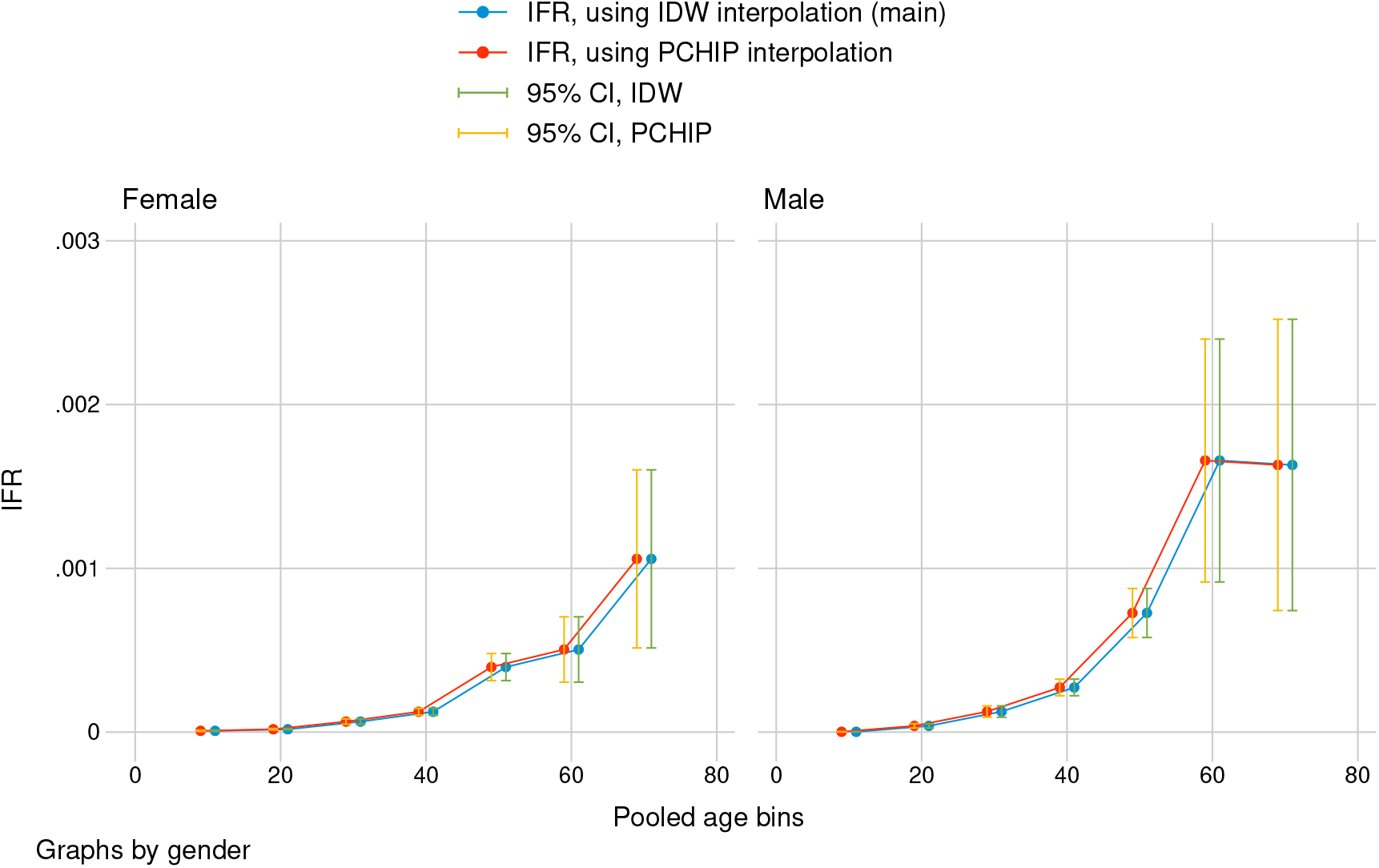
Karnataka: sensitivity analysis using piecewise cubic Hermite interpolation to estimate age bin share of deaths

Government reports provide age-shares of deaths in age bins of the form 11-20, 21-30, etc. To match seroprevalence estimates, we interpolate age-shares of deaths in the form 10-19, 20-29, etc. Main specification uses the inverse distance weighted average (IDW) to interpolate age shares. sensitivity analysis uses piecewise cubic Hermite interpolation. Interpolation was done with Stata package mipolate. Graphs by gender with 95% confidence intervals. Standard errors reflect propagation of error from design-based uncertainty of seroprevalence estimates. Confidence intervals are not reported for Bangalore due to small sample size, and age-specific estimated IFRs in Bangalore should not be interpreted as conclusive. IFRs are calculated in age bins 0-9, … 60-69, and 70+.

## 1 Supplementary Tables

**Table S1.** Zone-wise case multipliers for main and higher seroprevalence estimates based on sampled wards

| | | No. infections | | | $\gamma_z$ | |
| --- | --- | --- | --- | --- | --- | --- |
| *Ward* | *Zone* | *BMC report* | *main SP* | *high SP* | *main SP* | *high SP* |
| (1) | (2) | (3) | (4) | (5) | (6) | (7) |
| F North | City | 4,017 | 190,652 | 211,835 | 47.46 | 52.73 |
| M West | Eastern | 2,965 | 139,791 | 155,322 | 47.15 | 52.39 |
| R North | Western | 2,421 | 145,413 | 161,569 | 60.06 | 66.74 |

“Number of infections, main SP” refers to the estimated seroprevalence ( using the midpoint estimated sensitivity of the antibody test) multiplied by population in each sampled ward. “Number of infections, high SP” uses lowest bound sensitivity of the antibody test. Case multiplier “ $\gamma_z$ , main SP” (Column 6) was calculated by dividing Column 4 by Column 3. “ $\gamma_z$ , high SP” (Column 7) was calculated by dividing Column 5 by Column 3. Main SP indicates seroprevalence estimated from midpoint of two published estimates of sensitivity of the antibody test. High SP indicates seroprevalence was estimated using the minimum sensitivity and maximum specificity of the antibody test, generating a upper-bound estimate.

**Table S2.**
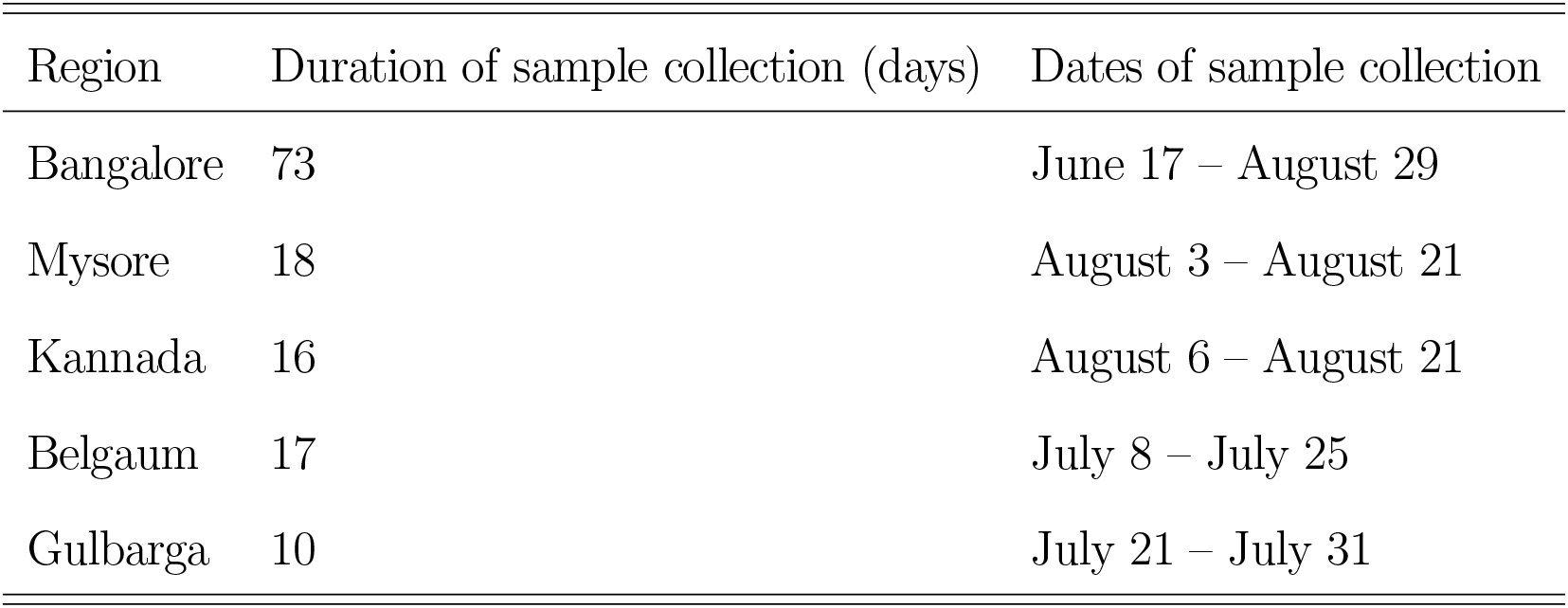
Karnataka: duration of sample collection by region

## Footnotes

1 In Belgaum, the target date was July 27th; we used July 28. In the sensitivity test, we used August 11 instead of August 10, which was unavailable.

## Notes

### Competing Interest Statement

The authors have declared no competing interest.

### Funding Statement

This paper was partially funded by Emergent Ventures Grant #466, given to A. Malani.

### Author Declarations

The research was not subject to an IRB/oversight body, because there was no interaction with human subjects and no privately identifiable information. Data gathered from bio-samples have been reported in separate papers, but aggregated and anonymized for use in this paper.

